# AFQuery: a bitmap-indexed, capture-aware allele frequency engine for clinical genomics cohorts

**DOI:** 10.64898/2026.05.15.26353174

**Authors:** Gabriel Santos-Díaz, Noemí Toro-Barrios, Rosario Carmona, Graciela Uría-Regojo, Rocío Jiménez-Arias, Xaquín Gurriarán, Paula Ramilo, Jorge Amigo, Pablo Mínguez, Joaquín Dopazo, Daniel López-López

**Affiliations:** Andalusian Platform for Computational Medicine, Andalusian Public Foundation Progress and Health (FPS), Sevilla, Spain; Computational Systems Medicine, Institute of Biomedicine of Seville (IBiS), Hospital Universitario Virgen del Rocío, Sevilla, Spain; Centre for Biomedical Network Research on Rare Diseases (CIBERER), Instituto de Salud Carlos III, Madrid, Spain; Department of Genetics & Genomics, Bioinformatics Unit, Instituto de Investigación Sanitaria-Fundación Jiménez Díaz University Hospital (IIS-FJD, UAM), Madrid, Spain; Fundación Pública Galega de Medicina Xenómica (FPGMX), SERGAS, Santiago de Compostela, Spain; Barcelonaβeta Brain Research Center (BBRC), Barcelona, Spain; Hospital del Mar Research Institute Barcelona (HMRIB), Barcelona, Spain

**Keywords:** allele frequency, variant classification, ACMG, bitmap index, capture kit, clinical genomics

## Abstract

**Motivation:** Allele frequency (AF) is central to clinical variant classification under ACMG/AMP guidelines. Public reference databases offer broad ancestry coverage, but local ancestries, rare-disease enrichment, and institutional case distributions are often underrepresented, so cohort-derived AF is a valuable complement. Computing accurate AF from institutional cohorts is nonetheless error-prone: even successive versions of the same capture kit cover substantially different target regions, and naive methods inflate the allele number (AN) at positions not shared by all kits, deflating AF and biasing ACMG frequency evidence toward pathogenic categories.

**Results:** We present AFQuery, a bitmap-indexed AF engine that computes capture-aware, ploidy-aware allele frequencies from pre-indexed Roaring Bitmaps in ∼ 14 ms per point query (∼34 ms for 1-Mbp region queries), independently of cohort size up to 50,000 samples. In simulated mixed-technology cohorts, capture-aware AN reduced AF mean absolute error 8–13-fold and removed the systematic bias toward pathogenic ACMG categories, yielding 10–45-fold fewer spurious pathogenic-evidence calls.

**Availability:** AFQuery is freely available under the MIT licence at https://github.com/babelomics/afquery.

---

Allele frequency (AF) is the most impactful evidence type in clinical variant classification, underpinning three ACMG/AMP criteria—BA1 (stand-alone benign), BS1 (benign strong), and PM2 (pathogenic moderate) (Richards et al., 2015). Population databases such as gnomAD (Karczewski et al., 2020) stratified by global ancestry, but they often under-represent local ancestries, rare-disease enrichment, and institutional case distributions; in one inherited-arrhythmia cohort, supplementing global databases with local cohort frequencies reclassified 32.2% of variants of uncertain significance (VUS) (Young et al., 2024). As clinical sequencing programmes mature, institutional variant databases accumulate samples from successive technology generations—panels, whole-exome sequencing (WES) with different capture kits, and whole-genome sequencing (WGS)— so accurate AF computation across heterogeneous cohorts becomes a prerequisite for maximising diagnostic yield.

Accurate AF estimation requires large sample sizes, making it tempting to query such heterogeneous cohorts as a whole. When cohorts combine WGS, WES with different capture kits, and targeted panels, however, AF computation becomes non-trivial. Standard methods compute allele number (AN) as twice the number of eligible samples at every position, ignoring that WES and panel samples lack coverage outside their capture regions. As a result, AN is inflated and the observed AF is correspondingly deflated. The problem in fact arises whenever any two technologies differ in target regions, including successive versions of the same vendor’s kit (Clark et al., 2011), an invisible source of AN inflation, since all samples are labelled “WES” regardless of kit version. Such kit-driven batch effects have been documented at scale (Wickland et al., 2021; Carson et al., 2014), yet no existing tool offers built-in AN computation on a per-position, per-technology basis. GQT (Layer et al., 2016) introduced bitmap-indexed genotype compression but lacks AF computation and metadata filtering; GEMINI (Paila et al., 2013) computes AF by linear SQLite scan without positional indexing; Hail (Hail Team, 2024) requires Spark infrastructure beyond most clinical settings; and BCFtools (Danecek et al., 2021) reprocesses all samples per query at *O*(*N*) cost without capture-aware AN. Joint-calling on gVCFs (e.g. GLnexus (Yun et al., 2020)) sidesteps the hom-ref/no-coverage ambiguity at source and is the recommended approach when gVCFs are available, but retrospective institutional databases often retain only variant-only VCFs.

We present AFQuery, a bitmap-indexed allele frequency engine for clinical genomics cohorts. AFQuery indexes genotype data from single-sample VCFs into Roaring Bitmaps (Lemire et al., 2018)—compressed bitsets stored in Apache Parquet files partitioned by chromosome. Because denser bitmaps compress more efficiently, query latency stays effectively constant as the cohort grows. Sample-level filters (sex, phenotype, technology) are precomputed as bitmaps too, so arbitrary subgroup queries reduce to bitmap intersection. The engine is file-based and serverless, running on standard workstations, HPC clusters, or containers without any database infrastructure. The genome build (GRCh37 or GRCh38) is fixed at database creation and determines pseudo-autosomal region boundaries for ploidy-aware AN computation on sex chromosomes.

AFQuery computes allele number per position, not per cohort. It first resolves the eligible sample set by intersecting precomputed metadata bitmaps; then, for each non-WGS technology, a capture index built from the technology’s BED file decides whether the queried position falls within its target regions. Each kit version is registered as a distinct technology with its own BED file, so a position targeted by SureSelect v6 but absent from v5 contributes only v6 samples to the AN denominator. AN is finally computed with ploidy awareness across autosomes and sex chromosomes (see Supplementary Methods §3 and Supplementary Table S1).

AFQuery supports four query modes: point queries for single variants, region queries for genomic intervals, batch queries for variant lists, and bulk export (`dump`) for complete allele frequency tables with optional stratification by sex, technology, or phenotype. The `annotate` command writes cohort allele frequencies back into VCF files in parallel, integrating cleanly with existing clinical pipelines. A complementary `variant-info` command returns the samples carrying a queried variant together with their metadata—sex, technology, phenotype codes, genotype and FILTER—supporting ACMG-style review without re-scanning the original VCFs.

Because AFQuery ingests variant-only VCFs rather than gVCFs, a sample’s absence at a given position does not by itself guarantee a confident reference-homozygous call. AFQuery addresses this with two layers of conservatism. First, BED-based exclusion structurally removes samples outside their technology’s capture regions from AN. Second, a dedicated `N_NO_COVERAGE` field— configurable via build- and query-time gates (`–min-pass, –min-observed, –min-dp, –min-gq, –min-quality-evidence`)—flags within-BED quality-failing samples that remain in AN but are not counted as hom-ref (Supplementary Methods §4). FILTER=PASS is always enforced at ingestion; non-PASS calls are retained in `fail_bitmap` for downstream reporting.

We evaluated AFQuery using chromosome 22 from the 1000 Genomes Project (1000 Genomes Project Consortium et al., 2015), scaling synthetically up to 50,000 samples. Point-query latency was 13.8 ms at 1,000 samples and 13.2 ms at 50,000 samples (warm cache, median over 50 replicates), confirming the expected constant-time behaviour (Figure 1A). Region queries (1 Mbp) completed in ∼34 ms and batch queries (100 variants) in ∼39 ms, again independently of cohort size; combined metadata filters (sex, phenotype, technology) added no measurable overhead. We benchmarked against BCFtools (Danecek et al., 2021), the de facto standard for VCF manipulation and AF computation. The two tools agreed closely: *R*^2^ *>* 0.99999 (MAE*<*10^−5^) over 1,106,181 common variants in 2,504 samples, with a single allele-count (AC) discrepancy attributable to multi-allelic representation and zero AN mismatches.

**Figure 1:**
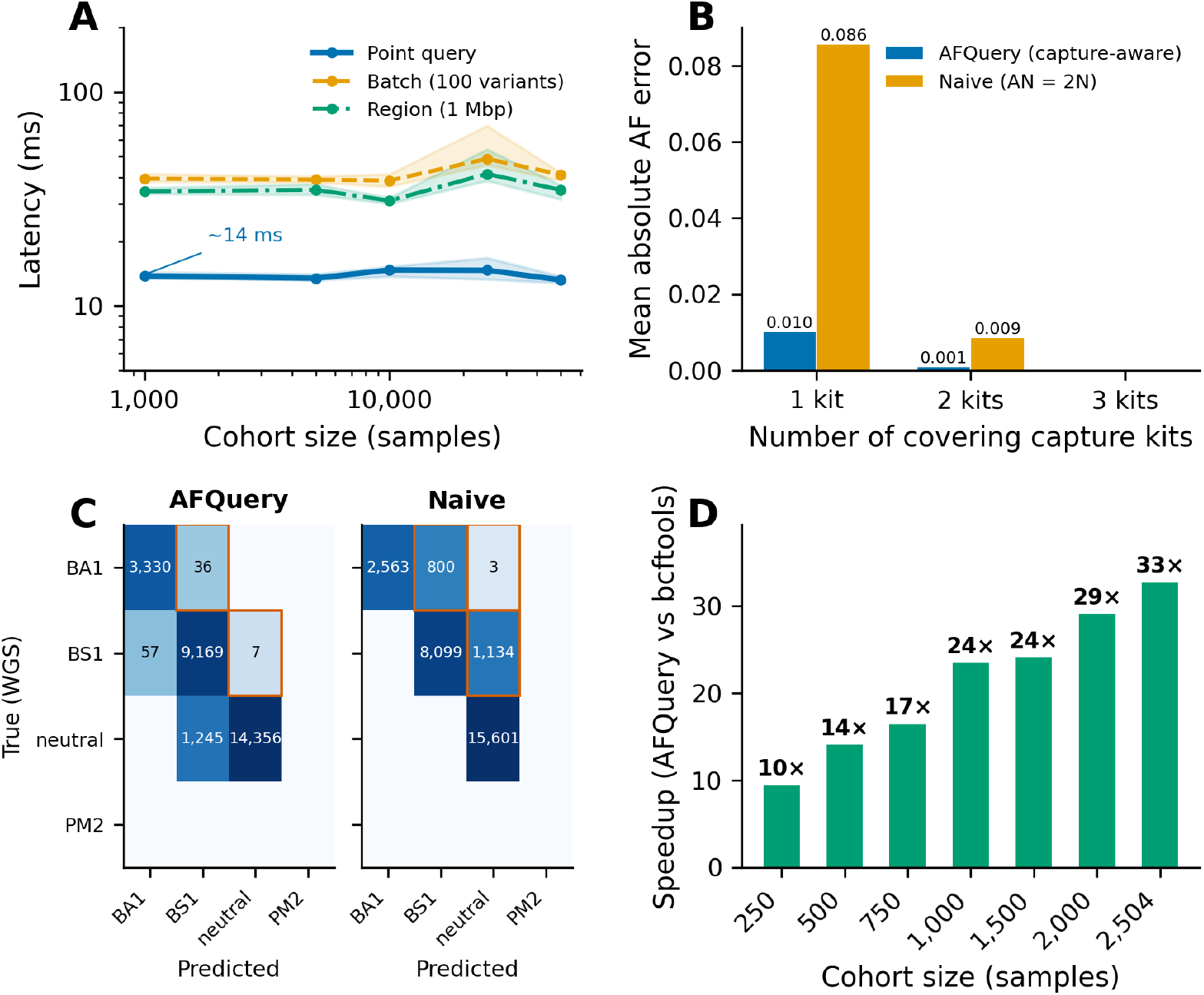
AFQuery performance and clinical accuracy. **(A)** Point-query latency stays essentially constant (∼14 ms) from 1,000 to 50,000 samples, illustrating the *O*(1) scaling afforded by Roaring Bitmap indexing; shaded areas: interquartile range (IQR) over 50 warm-cache replicates. **(B)** Mean absolute AF error stratified by the number of capture kits covering each position (extreme scenario: 800/150/50 across three Agilent SureSelect kits, chr22, 1,000 samples). AFQuery’s capture-aware AN eliminates error at three-kit positions and reduces it 8.4-fold at single-kit positions, where naive AN produces errors up to 0.086. **(C)** ACMG classification confusion matrices (cardiomyopathy thresholds: BA1 = 5%, BS1 = 0.1%, PM2 = 0.01%) for AFQuery (left) and naive (right) against WGS ground truth (28,200 chr22 variants, extreme scenario). Naive errors: 100% toward pathogenicity (1,937 variants, 6.87%); AFQuery residual errors: 97% toward benign (43 variants, 0.15%)—a 45-fold reduction. **(D)** Bulk-export speedup over BCFtools grows from 10× at 250 samples to 33 × at 2,504 samples (chr22 full dump). BCFtools scales linearly, whereas AFQuery’s per-sample cost decreases as bitmaps become denser. Dataset: chromosome 22, 1000 Genomes Project (1000 Genomes Project Consortium et al., 2015).

On bulk operations, AFQuery substantially outperformed BCFtools. Full chromosome 22 export completed in 7.0 s versus 3.8 min for BCFtools (2,504 samples)—a 33-fold speedup that grew with cohort size, from 10× at 250 samples to 33× at 2,504 across seven tested cohorts (Figure 1D). BCFtools `+fill-tags` is single-threaded by design and scales linearly with sample count (∼90 ms/sample), whereas AFQuery owes the observed speedup to two genuine architectural advantages: multithreading and Roaring Bitmap compression. For single point queries BCFtools was 1.6× faster (18 ms vs. 28 ms), reflecting AFQuery’s per-query DuckDB (Raasveldt and Mühleisen, 2019) connection overhead; this cost is amortised whenever consecutive queries reuse a single QueryEngine, as `annotate` does.

Database construction is a one-time cost that runs comfortably on commodity hardware: building from 10,000 single-sample VCFs (chromosome 22) completed in 26 s on 16 cores (1.5 GB peak memory), or in 127 s single-threaded. The resulting database occupied 13.5 MB versus 181.3 MB for the source VCFs (13.4× compression), and the ratio improves as cohorts grow. VCF annotation reached ∼25,000 variants/s on 4 cores, enough to annotate a typical 30,000-variant exome in about one second. Moreover, samples can be added or removed incrementally, without rebuilding the database from scratch.

To quantify how much target regions actually diverge between kit versions, we compared the chromosome 22 capture regions of three successive Agilent SureSelect Human All Exon kits (v5, v6, v7). Target sizes ranged from 1.08 to 1.37 Mbp—a 27% difference between versions of the same product line. Their union spanned 1.51 Mbp, of which only 57.3% was targeted by all three versions; the remaining 42.7% was covered by one or two kits only (Supplementary Table S8). Despite sharing a vendor and product name, nearly half of all targeted positions on chromosome 22 are not universally captured across three kit generations, leaving a large fraction of the exome exposed to AN inflation in any cohort that spans these versions.

We then evaluated capture-aware AN by assigning 1,000 samples to the three kits under three scenarios: balanced (334/333/333), skewed (600/300/100), and extreme (800/150/50, representing a partially-completed kit transition with a small legacy cohort). At three-kit positions, both methods produced zero AF error; at two- and single-kit positions, capture-aware computation reduced the mean absolute error (MAE) 8.4–13-fold across scenarios (Figure 1B; Supplementary Table S9). In the extreme scenario, the naive AF error at single-kit positions reached 0.95 (nearly the maximum possible deviation), whereas the capture-aware MAE stayed at 0.010. All error was confined to positions covered by fewer than three kits, and the bias was unidirectional: an inflated AN mechanically deflates AF (AC/AN). The effect is therefore structural, not an artefact of any particular random kit-assignment seed.

Applying the same ACMG frequency-based criteria with cardiomyopathy thresholds (BA1 = 5%, BS1 = 0.1%, PM2 = 0.01%) (Walsh et al., 2017) to the chromosome 22 variants covered in each scenario, we quantified shifts in the frequency-evidence category alone. In the extreme scenario (28,200 variants), naive AF produced 1,937 variants with toward-pathogenic shifts (6.87%) versus 43 for AFQuery (0.15%)—a 45-fold reduction (Figure 1C); reductions ranged from 10.6-fold (balanced) to 45-fold (extreme). Across all six tested settings (three kit distributions × two disease models), naive shifts were 100% toward pathogenicity, a deterministic consequence of the same AN-inflation mechanism, while AFQuery’s residual errors were 93–97% toward-benign, arising from sampling variance at positions covered by fewer technologies. All classification errors fell at positions covered by fewer than three kits. Because laboratories typically upgrade kits gradually, the skewed and extreme distributions are more representative than the balanced one, making capture-aware AN a critical requirement for reliable frequency-evidence assignment in cohorts that span multiple kit generations.

A few limitations should be noted. The capture-aware experiments demonstrate correct denominator bookkeeping under heterogeneous capture regions, but they rely on simulated technology assignments on chromosome 22; robustness to technology-specific calling differences awaits validation on cohorts with documented mixed-kit composition. The 50,000-sample scaling cohorts were generated by resampling with replacement from 2,504 real samples, so truly diverse biobank-scale data may exhibit different compression dynamics. WGS serves as ground truth despite known coverage heterogeneity in repetitive regions. Coverage eligibility is assessed per technology by default; per-sample resolution is nonetheless available by registering mosdepth-derived BEDs as one technology per sample (Supplementary §2). The ploidy model assumes binary sex and does not capture X-aneuploidies. Finally, the kit-overlap analysis covered chromosome 22, but the underlying mechanism—redesigned probe sets between versions—applies genome-wide.

## Data Availability

All data produced are available online at https://github.com/babelomics/afquery

https://babelomics.github.io/afquery

https://doi.org/10.5281/zenodo.19300230

## Data and code availability

AFQuery (MIT licence, Python ≥3.10; installable via pip, Bioconda, or Docker): https://github.com/babelomics/afquery; documentation: https://babelomics.github.io/afquery/. Benchmarking scripts, BED files, and result files at Zenodo (López-López, 2026). Input data: chromosome 22, 1000 Genomes Phase 3.

