## supplementary file for "AFQuery: a bitmap-indexed, capture-aware allele frequency engine for clinical genomics cohorts"

**5 References** **17**

**6 Supplementary Figures** **18**

### 1 Clinical motivation: local cohort AF beyond global databases

The value of ancestry- and cohort-matched allele frequencies is not theoretical. Recent reports across diverse populations and disease contexts have shown that incorporating local frequency data into ACMG/AMP-based interpretation substantively alters variant classification. Huang et al. [2026] combined whole-genome AF from 1,492 Taiwan Biobank individuals with a cohort of 802 inherited-retinal-degeneration probands, operationalising ACMG criterion PS4 in an ancestry-matched case–control framework; the approach upgraded multiple variants from VUS to Likely Pathogenic that global references alone had left unresolved. Agaoglu et al. [2024] likewise reported that the genomic disparity between Turkish breast-cancer patients and the populations underlying gnomAD shifts the classification of cancer-susceptibility-gene variants. Finally, Dawood et al. [2024] showed that variant-interpretation inequities in underrepresented populations can be partially mitigated by combining multiplexed functional data with ancestry-aware cohort frequencies.

These independent examples converge on a common requirement, and motivate the engineering choices behind AFQuery: a portable, capture-aware engine that lets clinical laboratories produce institutional AF tables in their own populations without depending on global aggregations alone.

### 2 Supplementary Methods

#### 2.1 Software Architecture and Data Model

AFQuery is a file-based allele frequency query engine implemented in Python (≥3.10) that indexes genotype data from single-sample VCFs into Roaring Bitmaps [Lemire et al., 2018] stored in Apache Parquet files. The system requires no server process or database daemon: all data resides in a directory structure readable by any process with filesystem access.

**Input data.** AFQuery accepts a manifest tab-separated values (TSV) file specifying one row per sample with columns: `sample_name`, `vcf_path`, `sex` (male or female), `tech_name` (sequencing technology identifier), and `phenotype_codes` (semicolon-separated free-text identifiers; institutions may use Human Phenotype Ontology (HPO), International Classification of Diseases, 10th revision (ICD-10), Mondo Disease Ontology (MONDO), or local codes interchangeably—AFQuery treats them as opaque strings and indexes each unique code as a precomputed bitmap). Each non-WGS technology must have an associated BED file defining its capture regions. The genome build (GRCh37 or GRCh38) is specified at database creation and determines pseudo-autosomal region (PAR) boundaries.

**On-disk layout.** The database directory contains: (1) Hive-partitioned Parquet files under `variants/{chrom}/bucket_{id}.parquet`, where each bucket spans 1 Mbp of genomic positions; (2) a SQLite metadata database (`metadata.sqlite`) storing sample information, technology definitions, phenotype associations (many-to-many), precomputed filter bitmaps, and a changelog; (3) capture index pickle files (`capture/tech_{id}.pickle`) for non-WGS technologies; and (4) a `manifest.json` recording build configuration, genome build, and schema version.

**Per-variant representation.** Each variant is identified by its position, reference allele and alternate allele, and is encoded as three core Roaring Bitmaps: `het_bitmap` for samples heterozygous with FILTER=PASS, `hom_bitmap` for samples homozygous alternate with FILTER=PASS, and `fail_bitmap` for samples with a non-PASS record at the site (either a carrier whose call failed a filter, or a sample with a missing genotype at a site whose FILTER≠PASS). Each bitmap uses dense integer sample identifiers as bit positions, so set operations—intersection, union, difference, cardinality—run in microseconds via the pyroaring library (≥0.4.8). Databases built with the coverage-quality filters (§2.4) carry two additional bitmaps, `filtered_bitmap` and `quality_pass_bitmap`, which support the `N_NO_COVERAGE` machinery.

**Precomputed filter bitmaps.** At database creation, sample bitmaps are precomputed and stored in SQLite for each sex category (male, female), each sequencing technology and each phenotype code. Arbitrary filter combinations can then be resolved at query time by intersecting the relevant precomputed bitmaps, a sub-microsecond operation that bypasses any scan of sample metadata.

#### 2.2 VCF Ingestion and Build Pipeline

**Ingestion phase.** Each single-sample VCF is parsed in a parallel worker (Python ProcessPoolExecutor) using cyvcf2 (≥0.30). For every variant, the worker extracts the genotype (allele count: 0, 1, or 2) and FILTER status and writes them to a temporary per-sample Parquet file with the schema `chrom`, `pos`, `ref`, `alt`, `gt_ac`, `sample_id`, `filter_pass`, `dp`, `gq`, `qual` (the last three populated when the corresponding `FORMAT` fields are present and feed the build-time quality gates described in §2.4).

**Consolidation phase.** DuckDB (≥0.10) [Raasveldt and Mühleisen, 2019] reads all temporary per-sample Parquet files, groups rows by (chrom, pos, ref, alt), and constructs the per-variant Roaring Bitmaps by collecting sample identifiers into het, hom, and fail sets. Samples with allele count 1 and FILTER=PASS are added to `het_bitmap`; allele count 2 and FILTER=PASS to `hom_bitmap`; non-PASS to `fail_bitmap`.

**Output.** The consolidated data is written as Hive-partitioned Parquet files, one per 1-Mbp bucket per chromosome. Bucketing at this granularity lets DuckDB's predicate pushdown read only the file that contains a queried position, so point queries never trigger a full-chromosome scan.

**Incremental updates.** Samples can be added or removed without rebuilding the entire database. The `update-db` command reprocesses only affected Parquet files and updates the metadata accordingly.

##### 2.3 Capture-Aware Allele Number Computation

AFQuery computes allele number (AN) per position rather than per cohort, so each queried position is normalised by the samples that actually carry sequencing coverage there.

**Capture index construction.** For each non-WGS technology, the associated BED file is loaded via PyRanges (≥0.1.2) [Stovner and Sætrom, 2020] and converted to per-chromosome sorted arrays of interval start and end positions. The resulting `CaptureIndex` object is serialised as a pickle file. Position coverage lookup uses binary search (`bisect.bisect_left`) with backward scan for overlapping intervals, yielding $O(\log n)$ per query. WGS technologies use a fast-path `CaptureIndex` with `always_covered=True` (constant-time).

**Two-stage eligibility model.** AN computation proceeds in two stages.

*Stage 1—Sample-level filtering.* The eligible sample set is obtained by intersecting precomputed bitmaps: eligible = phenotype_bm ∩ sex_bm ∩ tech_bm (when no filters are supplied, every sample is eligible).

*Stage 2—Position-level coverage.* For each non-WGS technology, the capture index is then queried to decide whether the position falls within its target regions; if so, that technology's sample bitmap is added to the covered set by union. The final eligible set is eligible_at_position = eligible ∩ covered, which guarantees that WES and panel samples contribute to AN only at positions inside their capture regions.

**Ploidy-aware AN formula.** Once the eligible set at a position is determined, AN is computed according to chromosome and sex:

Supplementary Table S1: Ploidy-aware allele number formulas by chromosome class.

| Chromosome class | AN formula |
|---|---|
| Autosomes (chr1–22) | $2 \times \lvert \text{eligible} \rvert$ |
| chrX PAR regions | $2 \times \lvert \text{eligible} \rvert$ |
| chrX non-PAR | $2 \times \lvert \text{eligible} \cap \text{female} \rvert + \lvert \text{eligible} \cap \text{male} \rvert$ |
| chrY | $\lvert \text{eligible} \cap \text{male} \rvert$ |
| chrM | $\lvert \text{eligible} \rvert$ |

PAR boundaries follow established coordinates: GRCh38 chrX PAR1 [10,001–2,781,479], PAR2 [155,701,383–156,030,895]; GRCh37 chrX PAR1 [60,001–2,699,520], PAR2 [154,931,044–155,260,560].

**Ploidy-aware AC formula.** The complementary AC computation splits the eligible set into haploid and diploid subsets (for chrX non-PAR, for example, males are haploid and females diploid) and takes the form

$$\text{AC} = |\text{carriers} \cap \text{haploid}| + |\text{het} \cap \text{diploid}| + 2 \times |\text{hom} \cap \text{diploid}|,$$

with carriers = het ∪ hom. Allele frequency follows as AF = AC/AN whenever AN $> 0$; positions with AN $= 0$ are omitted from the output.

#### 2.4 Coverage-Evidence Model and the `N_NO_COVERAGE` Field

AFQuery ingests single-sample variant-only VCFs rather than gVCFs. Variant-only VCFs do not record reference-homozygous calls, so the absence of an entry at a position is intrinsically ambiguous: it can mean either that the sample was sequenced and is genuinely homozygous reference, or that the sample was not sequenced deeply enough to support a confident hom-ref call. Resolving this ambiguity correctly is essential for accurate allele-frequency computation in mixed-technology cohorts.

**Fully-covered vs. partially-covered technologies.** A technology registered without a BED file (typically WGS) is treated as *fully-covered*: every position is assumed sequenced, so a sample with no entry there is recorded as hom-ref. A technology registered with a BED file (whole-exome kits, gene panels) is instead *partially-covered*: the BED guarantees that the region was *targeted* by the assay, but not that any particular sample was sequenced deeply enough there to support a confident hom-ref call.

**The `N_NO_COVERAGE` field.** To label the uncertain non-carrier subset without inflating allele frequency, AFQuery emits a per-position counter `N_NO_COVERAGE`. The genotype invariant for the eligible set at a queried position is

$$\text{N\_HET} + \text{N\_HOM\_ALT} + \text{N\_HOM\_REF} + \text{N\_FAIL} + \text{N\_NO\_COVERAGE} = n_{\text{eligible}}.$$

Samples in `N_NO_COVERAGE` remain in `eligible` and therefore continue to contribute to AN (analogously to `N_FAIL`), so allele frequency stays conservative—the field never inflates AF. Two structural rules hold by construction: carriers (`het`, `hom`, or `fail` calls) are never reclassified, and samples on fully-covered technologies are never gated.

**Cohort-evidence gates (query-time, no rebuild).** Two complementary flags decide, on a per-position basis, whether each partially-covered technology has shown enough cohort-level evidence to trust a hom-ref call. The first, `--min-pass K`, requires at least $K$ PASS carriers (het ∪ hom) for that technology at the position; the second, `--min-observed K`, instead counts *any* recorded carrier, including FILTER-failing calls (het ∪ hom ∪ fail). Whenever a technology falls short of the active threshold, all of its non-carrier samples at that position are reassigned from `N_HOM_REF` to `N_NO_COVERAGE`. If both flags are set to non-zero values, both conditions must be met simultaneously.

**Quality-aware filtering (build-time + query-time).** When the input VCFs carry usable read depth (`FORMAT/DP`), genotype quality (`FORMAT/GQ`) or `QUAL` fields, the `create-db` command can apply additional thresholds at build time: `--min-dp`, `--min-gq`, `--min-qual` and `--min-covered K`. A carrier counts as quality-passing only when every active threshold is satisfied, and a position is regarded as "trusted" for a given partially-covered technology only if at least $K$ such quality-passing carriers are present. The selected thresholds are stored alongside the database and re-applied automatically by `update-db --add-samples`. A complementary query-time flag, `--min-quality-evidence K`, can tighten the gate without rebuilding, provided the database was built with the relevant quality data.

**Composition.** Taken together, the gates described above partition the eligible set so that `N_NO_COVERAGE` is the union of three subsets: samples whose technology failed the build-time `--min-covered` gate, samples whose technology failed `--min-pass` or `--min-observed` at query time, and samples whose technology failed `--min-quality-evidence`. Carriers are never included and no sample is counted twice. Worked examples and recommended threshold profiles are provided at https://babelomics.github.io/afquery/latest/advanced/coverage-evidence/.

#### 2.5 Query Execution

**Point queries.** For a single variant, the engine resolves the Parquet bucket file (bucket_id = pos // 1,000,000), executes a DuckDB SQL query with predicate pushdown, deserialises the three bitmaps, intersects them with the eligible bitmap, and computes AC/AN/AF. A new DuckDB connection is created per query (~5 ms overhead) to ensure thread safety.

**Batch queries.** For lists of variants on the same chromosome, AFQuery selects between two SQL strategies depending on batch size. Batches below 10,000 positions are issued with an IN clause and parameter binding; from 10,000 positions onwards, the engine switches to a temporary-table join to sidestep DuckDB's IN-clause performance degradation at scale. The eligible set and AN are computed once per unique position and cached, so variants that share a position do not pay the cost twice.

**Region queries.** For genomic intervals, a range predicate (`WHERE pos BETWEEN ? AND ?`) is applied across all bucket files in the interval. Results are sorted by (pos, alt).

**Sample-level retrieval (`variant-info`).** The complementary `variant-info` subcommand returns the list of samples carrying a queried variant together with their metadata (sex, technology, phenotype codes, genotype and FILTER status). It accepts the same metadata filters as `query` and supports text, TSV and JSON outputs, making it a direct follow-up to any `query` result for ACMG-style review without forcing the user to re-scan the original VCFs.

#### 2.6 VCF Annotation and Bulk Export

**Annotation.** The `annotate` command adds cohort allele frequencies to a VCF file through a three-phase pipeline. First, the *collect* phase buffers the input variants and groups them by (chromosome, bucket_id). Next, the *compute* phase dispatches the buffered groups to parallel workers (ProcessPoolExecutor, with one QueryEngine per subprocess), which compute AC/AN/AF per variant. Finally, the *write* phase emits the results in the original VCF order. Eight INFO fields are added: AFQUERY_AC, AFQUERY_AN, AFQUERY_AF, AFQUERY_N_HET, AFQUERY_N_HOM_ALT, AFQUERY_N_HOM_REF, AFQUERY_N_FAIL and AFQUERY_N_NO_COVERAGE. Multi-allelic sites are handled per ALT allele, with Number=A for AC, AF and the genotype counts, and Number=1 for AN and N_FAIL.

**Bulk export.** The `dump` command generates complete allele frequency tables, parallelised by (chromosome, bucket) work units. Optional stratification produces separate AC/AN/AF columns for each combination of sex, technology, and phenotype categories.

#### 2.7 Benchmarking Datasets

**Base dataset.** All benchmarks used chromosome 22 from the 1000 Genomes Project Phase 3 [1000 Genomes Project Consortium et al., 2015] (2,504 samples), split into single-sample VCFs.

**Synthetic scaling.** To evaluate performance beyond the available 2,504 real samples, cohorts of 1,000, 5,000, 10,000, 25,000, and 50,000 samples were created by resampling with replacement from real genotypes (random seed = 42). This design tests bitmap operations under realistic AF distributions but introduces duplicate genotype patterns that may overstate compression efficiency relative to truly diverse cohorts; the constant-time claim should therefore be interpreted as cohort-size scaling under the available data, not as direct generalisation to biobank-scale cohorts of unique individuals.

**BCFtools comparison subsets.** To evaluate how performance scales with real (non-synthetic) cohort size, seven subsets of 250, 500, 750, 1,000, 1,500, 2,000 and 2,504 samples were drawn from the 1000 Genomes Phase 3 data. Three operations were benchmarked at each size: single-variant point query, subset query (filtering by sex) and full chromosome dump (all variants with allele frequencies).

With these seven sizes the scaling behaviour of each tool becomes apparent: BCFtools is linear in sample count (~90 ms/sample for the dump operation), whereas AFQuery scales sublinearly because Roaring Bitmap compression becomes increasingly efficient on denser bitsets. Note that BCFtools `+fill-tags` is single-threaded by design, so the reported speedups combine the benefit of parallelism with that of bitmap indexing—both legitimate operational advantages of the AFQuery architecture.

**Capture kit simulation.** To evaluate capture-aware AN computation, 1,000 samples were assigned to three Agilent SureSelect capture kits (v5, v6, v7) in three scenarios: balanced (334/333/333), skewed (600/300/100), and extreme (800/150/50). A WGS ground truth database (all samples, full coverage) provided reference allele frequencies. Naive allele frequencies were computed from the same database by ignoring capture regions (AN $= 2 \times N_{\text{eligible}}$ at all positions).

#### 2.8 Benchmarking Hardware and Software

All benchmarks were executed on a single node of an HPC cluster with Lustre parallel filesystem storage.

All dependencies are pinned in the benchmark conda environment specification (`benchmarks/envs/benchmark.yaml`).

#### 2.9 Statistical Methods

**Performance metrics.** Median latency (ms) with interquartile range (Q1–Q3) was reported for all timing benchmarks. Replicates: 50 (warm-cache query latency, after 3 discarded warmup iterations), 10 (BCFtools comparison), 3 (build and annotation). Cold-cache queries measured first-access overhead before filesystem caching.

Supplementary Table S2: Hardware specifications.

| Component | Specification |
|---|---|
| CPU | Intel Xeon Gold 6230R @ 2.10 GHz (52 cores) |
| RAM | 370 GB |
| OS | Rocky Linux 8.5 (kernel 4.18.0-348.12.2.el8_5.x86_64) |
| Storage | Lustre parallel filesystem |

Supplementary Table S3: Software versions.

| Software | Version |
|---|---|
| AFQuery | v0.3.2 |
| Python | 3.13.12 |
| DuckDB | 1.5.1 |
| pyroaring | 1.0.4 |
| pyarrow | 23.0.1 |
| bcftools | 1.23.1 |
| cyvcf2 | 0.32.1 |
| Snakemake | 9.18.2 |

**AF accuracy.** Mean absolute error (MAE) was computed between test and ground-truth (WGS) allele frequencies, overall and stratified by the number of capture kits covering each position (1, 2, or 3 kits). AN inflation ratio was computed per variant as $AN_{naive}/AN_{afquery}$.

**ACMG classification.** Two disease-specific threshold sets were applied: cardiomyopathy (BA1 = 5%, BS1 = 0.1%, PM2 = 0.01%) as recommended for sarcomeric cardiomyopathy by Walsh et al. [2017] and Whiffin et al. [2017], and a stricter illustrative metabolic-disease profile (BS1 = 0.01%, PM2 = absent). Variants were classified as BA1 (AF $\geq$ BA1), BS1 (AF $\geq$ BS1), PM2 (AF $\leq$ PM2), or neutral. Each variant's classification under AFQuery and naive AN was compared against WGS ground truth. Discordances were classified as toward-pathogenic (shift from benign to less-benign or pathogenic-supporting category) or toward-benign (reverse). BS1 recall was computed as the fraction of true BS1 variants correctly classified.

**Concordance.** $R^2$ was computed over 1,106,181 common variants on chromosome 22 (2,504 samples) between AFQuery and BCFtools allele frequencies. Per-variant AC and AN mismatches were counted.

#### 2.10 Data and Code Availability

AFQuery source code is available at `https://github.com/babelomics/afquery` under the MIT licence. The package can be installed via `pip install afquery`, Bioconda, or Docker. Documentation is available at `https://babelomics.github.io/afquery/`.

Benchmarking scripts and result files are deposited at Zenodo [López-López, 2026]: `https://doi.org/10.5281/zenodo.19300230`. The full benchmark suite (four performance experiments—query latency scaling, build performance, annotation throughput, AFQuery vs. BCFtools—and the capture-kit ACMG analysis) is orchestrated by a unified Snakemake pipeline and is fully reproducible:

```
cd benchmarks && snakemake --cores 32 --use-conda performance_all capture_kit_all
```

### 3 Supplementary Use Case: Per-sample Coverage via mosdepth

When per-sample binary alignment map (BAM) files are available but gVCFs have not been generated, AFQuery can still incorporate per-sample coverage information. The approach is to register each sample as a unique "technology" whose BED file is derived from its own coverage profile. This refines the eligibility model from per-technology to per-sample granularity, at the cost of a technology table that grows linearly with the cohort.

#### 3.1 Objective

To compute allele frequencies that account for per-sample coverage rather than assuming that every position in a kit-level BED is covered in every sample sequenced with that kit. A user-defined minimum coverage threshold determines which genomic positions are eligible for each individual sample, so the reference-homozygote count at a variant is normalised over the subset of samples that truly cover that position.

#### 3.2 Scope

This protocol applies to datasets generated from a complete DNA-seq pipeline that produces aligned BAM files and variant-only VCFs. All samples should be processed with the same reference genome, software versions, and parameter configurations to ensure comparability of allele frequency estimates. The protocol complements (rather than replaces) the standard per-technology workflow described in Supplementary Methods §2.3; if gVCFs are available, computing AF directly from them is simpler and is the recommended approach.

#### 3.3 Requirements

- **Software:** mosdepth [Pedersen and Quinlan, 2018] (https://github.com/brentp/mosdepth) and AFQuery (≥v0.3.2).
- **Input data:** per-sample aligned BAM files; corresponding single-sample variant-only VCFs; the reference genome (e.g. GRCh38); a manifest TSV in AFQuery format.
- **Storage:** one BED file per sample plus the standard AFQuery database directory.

#### 3.4 Procedure

**Step 1 — Generate per-sample coverage BED.** For each sample, run mosdepth with a minimum-coverage quantisation (default ≥10×). The output prefix passed to mosdepth determines the resulting filename; in the example below we use BED10X_<sample_name>:

```
mosdepth --quantize 10: -n -x ${output_prefix} ${bamfile}
```

The output contains a `quantized.bed.gz` with four columns: chromosome, start, end, and coverage label (`10:inf` for regions with depth ≥10×). The coverage threshold can be adjusted via `--quantize` (e.g. `5:` for ≥5×).

**Step 2 — Reduce to a 3-column BED.** AFQuery's capture index loader expects standard BED (chrom, start, end). Drop the fourth column:

```
zcat ${output_prefix}.quantized.bed.gz \
  | awk 'BEGIN{OFS="\t"} {print $1, $2, $3}' \
  > ${output_prefix}.bed
```

**Step 3 — Prepare the manifest.** Create a manifest TSV following the standard AFQuery schema (`sample_name`, `vcf_path`, `sex`, `tech_name`, `phenotype_codes`). The `tech_name` column carries the per-sample technology label that links each sample to its BED file. Use the BED file's basename (without extension): `tech_name = BED10X_<sample_name>`.

**Step 4 — Organise BED files.** Place every per-sample BED file into a single directory and pass it to `create-db` with `--bed-dir`:

```
afquery create-db \
  --manifest samples.tsv \
  --output-dir ./db/ \
  --genome-build GRCh38 \
  --bed-dir ./bed_dir/
```

AFQuery loads one capture index per technology (and therefore one per sample), and the two-stage eligibility model (Supplementary Methods §2.3) operates at sample granularity automatically—no further configuration is required at query time.

##### 3.5 Caveats and Trade-offs

- **When to use this workflow.** The recipe is designed for the case in which BAMs are available but gVCFs have not been generated. Whenever gVCFs already exist, computing AF directly from them is simpler and yields equivalent or better fidelity, and should be preferred.
- **Scale.** Per-sample technologies make the technology table grow linearly with the cohort. AFQuery's capture-index lookup is $O(\log n)$ per BED, so query latency remains constant in practice up to the scales reported in this study; database size grows with the number and complexity of per-sample BEDs.
- **Threshold choice.** A 10× threshold is a reasonable default for clinical exome and panel data; for low-coverage or targeted assays, lower thresholds (5×) may be appropriate. The same threshold should be applied uniformly across the cohort to preserve comparability.
- **Interaction with `N_NO_COVERAGE`.** Per-sample BEDs make the per-technology hom-ref ambiguity less acute because each sample's eligibility is decided by its own coverage data. The cohort-evidence and quality-aware gates documented in Supplementary Methods §2.4 remain available as a complementary layer of filtering.

### 4 Supplementary Results

#### 4.1 Query Performance Scaling

We evaluated AFQuery query latency across cohort sizes from 1,000 to 50,000 samples on chromosome 22, using synthetic cohorts generated by resampling from the 1000 Genomes Project Phase 3.

Point query latency (warm cache, median over 50 replicates) was 13.81 ms at 1,000 samples and 13.23 ms at 50,000 samples—effectively constant, demonstrating $O(1)$ scaling with cohort size (Supplementary Table S4). Cold-cache queries showed marginally higher latency (14.71 ms at 1,000 samples, 16.60 ms at 50,000 samples) due to initial filesystem I/O overhead.

Region queries (1 Mbp window) completed in ∼34 ms, and batch queries in ∼39 ms (100 variants) and ∼133 ms (1,000 variants), all constant across cohort sizes. Combined metadata filters (sex + phenotype + technology) produced point query latency in the range 13–16 ms with no significant overhead compared to unfiltered queries.

Supplementary Table S4: Query latency by type and cohort size (ms, median).

| Query type | 1,000 | 5,000 | 10,000 | 25,000 | 50,000 |
|---|---|---|---|---|---|
| Point (warm) | 13.81 | 13.47 | 14.72 | 14.68 | 13.23 |
| Point (cold) | 14.71 | 14.42 | 15.18 | 16.42 | 16.60 |
| Region (1 Mbp) | 34.38 | 34.90 | 31.05 | 41.42 | 34.84 |
| Batch (100) | 39.36 | 38.92 | 38.58 | 48.73 | 41.13 |
| Batch (1,000) | 132.88 | 133.06 | 118.64 | 161.22 | 132.28 |
| Combined filters | 16.09 | 15.21 | 13.40 | 13.98 | 13.22 |

#### 4.2 Build Performance

Database construction time scaled approximately linearly with cohort size and benefited from thread-level parallelism (Supplementary Figure S1). At 32 threads, chromosome 22 build times were 4.81 s (1,000 samples), 12.35 s (5,000 samples), and 24.25 s (10,000 samples). Single-thread build times were 20.54 s, 63.96 s, and 127.34 s respectively, yielding thread speedups of 4.3× (1,000 samples) and 5.2× (5,000 and 10,000 samples).

Per-sample build cost decreased with cohort size: from 20.5 ms/sample (1,000 samples, single-thread) to 12.7 ms/sample (10,000 samples), a 38% reduction attributable to amortisation of DuckDB consolidation overhead across more samples. At 32 threads, the reduction was more pronounced: from 4.8 ms/sample (1,000) to 2.4 ms/sample (10,000), a 50% improvement.

Peak resident set size (RSS) at 32 threads was 300.6 MB for 1,000 samples and 1,538.7 MB (1.5 GB) for 10,000 samples.

Supplementary Table S5: Build performance (chr22).

| Samples | 1 thread (s) | 4t | 8t | 16t | 32t | Speedup | Peak RSS |
|---|---|---|---|---|---|---|---|
| 1,000 | 20.54 | 6.38 | 5.63 | 4.91 | 4.81 | 4.3× | 300.6 MB |
| 5,000 | 63.96 | 19.51 | 16.59 | 12.36 | 12.35 | 5.2× | 871.3 MB |
| 10,000 | 127.34 | 50.28 | 37.72 | 26.18 | 24.25 | 5.2× | 1,538.7 MB |

Peak RSS scales with thread count and storage I/O latency. The `create-db` default working-memory budget (`--build-memory 2GB`) is sufficient for the configurations above on Lustre storage;

on slower storage or larger cohorts the budget may need to be raised (`--build-memory 4GB`).

The build phase decomposes into *ingest* (per-sample VCF parsing and per-sample Parquet generation) and *consolidate + build* (DuckDB grouping, Roaring Bitmap construction, partitioned Parquet output); these phases scale differently with thread count, with consolidate saturating earlier (close to its asymptote at 16 threads on our Lustre setup, with marginal improvement at 32), whereas ingest—dominated by per-sample VCF parsing—continues to benefit from additional workers. A per-phase wall-clock breakdown is not summarised here as it depends strongly on storage subsystem latency.

Independent measurements on alternative HPC infrastructure (faster storage, identical 32-core configuration) report build speedups up to $\sim$10× and ingest speedups up to $\sim$30× at 32 workers. The final reported values depend on the storage subsystem; the configurations above reflect our Lustre-based setup. For everyday operation on commodity hardware ($\leq$16 cores), the 8- to 16-thread region offers the best compromise between throughput and resource usage.

#### 4.3 VCF Annotation Throughput

Annotation throughput (variants/s) increased with both the number of input variants and the degree of thread parallelism (Supplementary Figure S2). At 32 threads, throughput was 17,203 variants/s for 10,000 input variants, 20,086 variants/s for 50,000 variants, and 30,460 variants/s for 100,000 variants. The increasing throughput with variant count reflects better amortisation of per-worker QueryEngine initialisation overhead.

At the maximum observed throughput (30,460 variants/s at 32 threads), a typical 30,000-variant exome annotates in approximately 1 second, and a 4,000,000-variant genome in approximately 2.2 minutes.

Supplementary Table S6: Annotation throughput (variants/s).

| Input variants | 1 thread | 4 threads | 32 threads | Speedup |
|---|---:|---:|---:|---:|
| 10,000 | 14,334 | 13,414 | 17,203 | 1.2× |
| 50,000 | 12,569 | 16,159 | 20,086 | 1.6× |
| 100,000 | 12,049 | 25,357 | 30,460 | 2.5× |

#### 4.4 AF Concordance with BCFtools

AFQuery allele frequencies were compared against BCFtools (`bcftools +fill-tags`) on chromosome 22 of the 1000 Genomes Project (2,504 samples). Over 1,106,181 common variants, $R^2 > 0.99999$ (Supplementary Figure S3), confirming near-exact AF concordance.

A closer look at the discordances showed a single variant (22:17996285 A>ATCTC) with an AC mismatch (AFQuery AC = 3,452 vs BCFtools AC = 3,444, $\Delta = 8$) and zero variants with AN mismatches. The discrepancy traces back to the multi-sample VCF, which contains two records at this position for the same alternate allele: a biallelic record (A>ATCTC; AC = 12) and a multi-allelic record (A>ATCTC,C; AC = 3,444). Eight of the samples are heterozygous for ATCTC in the biallelic record only; in the multi-allelic record they are homozygous for the other alternate (C). Because BCFtools computes AC line by line and treats each record independently, it retains only the multi-allelic record's count (3,444). AFQuery, on the other hand, ingests single-sample VCFs in which both records contribute genotypes for the same allele, so it picks up the eight additional carriers and reports the more complete count (3,452). The case therefore illustrates a structural advantage of per-sample ingestion: duplicate representations of the same allele are unified naturally, without requiring prior VCF normalisation.

An additional 4,056 variants were present only in BCFtools output; all had AC = 0 (reference-only entries in the multi-sample VCF), which AFQuery excludes by default.

##### 4.5 Storage Footprint

AFQuery databases were substantially smaller than the corresponding bgzipped single-sample VCFs (Supplementary Figure S4). Compression ratios improved with cohort size, reflecting more efficient Roaring Bitmap compression of denser bitsets:

Supplementary Table S7: Storage footprint (chr22).

| Samples | Raw VCFs | AFQuery DB | Ratio |
|---|---|---|---|
| 1,000 | 18.1 MB | 1.9 MB | 9.5× |
| 5,000 | 90.6 MB | 7.1 MB | 12.8× |
| 10,000 | 181.3 MB | 13.5 MB | 13.4× |

##### 4.6 SureSelect Kit Version Overlap (chr22)

To quantify target region divergence between successive Agilent SureSelect Human All Exon kits, we compared their chromosome 22 capture regions (Supplementary Table S8). Target sizes ranged from 1,079,651 bp (v7, 4,598 regions) to 1,366,356 bp (v6, 5,479 regions)—a 27% difference between versions of the same product line. Their union spanned 1,506,718 bp, of which only 863,486 bp (57.3%) were targeted by all three versions. The remaining 643,232 bp (42.7%) were covered by one or two kits only. Target region asymmetry was pronounced: v6 contributed 201,886 bp of unique target not present in either v5 or v7, and consequently only 63.2% of v6 targets fell within the three-way core, compared with 78.9% for v5 and 80.0% for v7.

Supplementary Table S8: Agilent SureSelect All Exon kit overlap (chr22).

| Quantity | v5 | v6 | v7 |
|---|---|---|---|
| Target size (bp) | — | 1,366,356 | 1,079,651 |
| Number of regions | — | 5,479 | 4,598 |
| % within 3-way core | 78.9 | 63.2 | 80.0 |

| Aggregate (chr22) | bp |
|---|---|
| Union of v5, v6, v7 | 1,506,718 |
| Shared by all three (3-way core) | 863,486 (57.3%) |
| Covered by one or two kits only | 643,232 (42.7%) |
| v6 unique target (not in v5 or v7) | 201,886 |

The 27% size difference and 42.7% non-shared target fraction observed across SureSelect v5/v6/v7 are consistent with independent comparisons of capture probe sets: Belova et al. [2022] reported substantial probe-set redesign between SureSelect v7 and v8, and Chilamakuri et al. [2014] documented broad target divergence across four commercial exome capture systems (Agilent, Illumina, Nimblegen, Roche). The kit-overlap quantification here therefore exemplifies a vendor-independent pattern, supporting the generalisability of the capture-aware AN argument beyond the SureSelect product line analysed.

##### 4.7 Capture-Aware AN: AF Error Across All Scenarios

We evaluated AF error across all three capture kit scenarios (Supplementary Figure S5). In all scenarios, capture-aware AN substantially reduced mean absolute AF error compared with naive computation:

Supplementary Table S9: Overall AF MAE by scenario.

| Scenario (kit split) | AFQuery MAE | Naive MAE | Reduction |
|---|---|---|---|
| Balanced (334/333/333) | 0.000760 | 0.009879 | 13.0× |
| Skewed (600/300/100) | 0.000872 | 0.009601 | 11.0× |
| Extreme (800/150/50) | 0.001323 | 0.011064 | 8.4× |

Error was exclusively concentrated at positions covered by fewer than three capture kits (10.8–16% of all covered variants). At positions covered by all three kits, both methods produced identical AF values (MAE = 0). The error reduction was most dramatic at single-kit positions, where naive MAE ranged from 0.043 (balanced) to 0.086 (extreme), while AFQuery MAE ranged from 0.003 to 0.010.

**AN inflation.** The maximum empirical AN inflation ratio ($AN_{naive}/AN_{afquery}$) at single-kit positions was 2.0× (balanced), 9.0× (skewed), and 19.0× (extreme). The balanced scenario's empirical maximum of 2.0× is below the theoretical maximum of 3.0× (all three kits / one kit covering) because not all three kits have identical sample counts and coverage boundaries. In the extreme scenario, the theoretical maximum of 20.0× (1,000 / 50 samples) is nearly reached.

##### 4.8 ACMG Classification Impact Across All Scenarios and Disease Models

ACMG frequency-based criteria were applied to the covered chromosome 22 variants in each scenario (29,662 balanced, 29,222 skewed, 28,200 extreme) using two disease-specific threshold sets (Supplementary Figure S6).

###### 4.8.1 Cardiomyopathy (BA1 = 5%, BS1 = 0.1%, PM2 = 0.01%)

Supplementary Table S10: Toward-pathogenic ACMG errors (cardiomyopathy).

| Scenario | AFQuery | Naive | Reduction | Naive toward-benign |
|---|---|---|---|---|
| Balanced | 159 (0.54%) | 1,688 (5.69%) | 10.6× | 0 |
| Skewed | 53 (0.18%) | 1,729 (5.92%) | 32.6× | 0 |
| Extreme | 43 (0.15%) | 1,937 (6.87%) | 45.0× | 0 |

**Error directionality.** In all three scenarios, naive errors were 100% toward-pathogenic (zero toward-benign errors). AFQuery residual errors were predominantly toward-benign: 92.5% (balanced), 96.2% (skewed), 97.3% (extreme)—a clinically conservative error profile.

**BS1 recall.** AFQuery correctly classified >98% of true BS1 variants in all scenarios (98.4%, 99.5%, 99.3% for balanced, skewed, extreme respectively), compared with ∼88% for naive computation (87.5%, 88.0%, 87.7%).

**Dominant error mode.** The most frequent naive error was BS1 → neutral (1,251 variants in the balanced scenario): variants that are truly benign-strong lost their BS1 classification because AN inflation deflated AF below the BS1 threshold. The second most frequent error was BA1 → BS1: variants exceeding the stand-alone benign threshold were downgraded to merely benign-strong.

###### 4.8.2 Metabolic Disease (BS1 = 0.01%, PM2 = absent)

Supplementary Table S11: Toward-pathogenic ACMG errors (metabolic disease).

| Scenario | AFQuery | Naive | Reduction | Naive toward-benign |
|---|---|---|---|---|
| Balanced | 23 (0.08%) | 437 (1.47%) | 19.0× | 0 |
| Skewed | 39 (0.13%) | 571 (1.95%) | 14.6× | 0 |
| Extreme | 36 (0.13%) | 803 (2.85%) | 22.3× | 0 |

The same directional pattern held: naive errors were 100% toward-pathogenic in all metabolic scenarios. The lower absolute error counts relative to cardiomyopathy reflect the stricter BS1 threshold (0.01% vs 0.1%), which is exceeded by fewer variants.

###### 4.8.3 Mechanism: Systematic Unidirectional Bias

The 100% toward-pathogenic directionality of the naive computation is structural, not coincidental. Inflating AN always increases the denominator of the AF fraction (AC/AN), and therefore always decreases AF. A lower AF can only shift a variant from benign categories toward pathogenic ones (BA1 → BS1, BS1 → neutral, neutral → PM2), never in the reverse direction. This is precisely why no toward-benign errors were observed in any of the six tested scenarios.

AFQuery's capture-aware AN removes that systematic bias by computing AN from the actual sequencing coverage at each position. Its residual errors (93–97% toward-benign) arise from an entirely different mechanism: at positions covered by only one or two kits, the reduced eligible sample count increases sampling variance, so AF estimates occasionally exceed the true population AF and push a neutral variant above the BS1 threshold.

### 6 Supplementary Figures

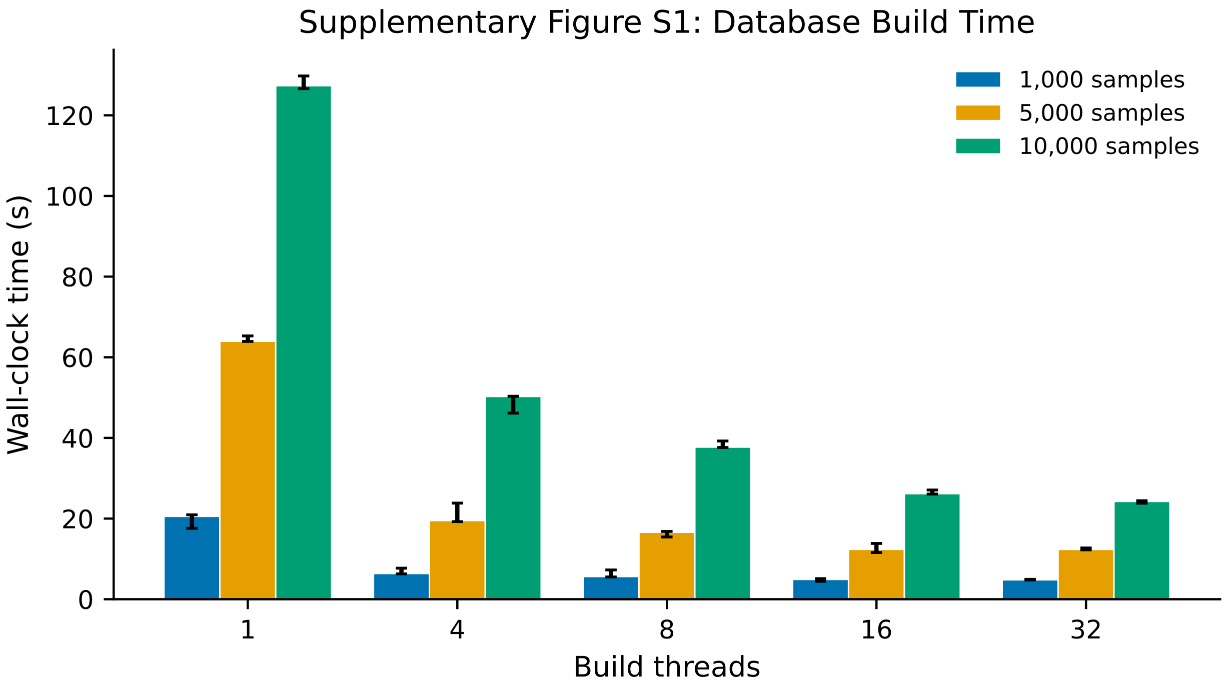

Supplementary Figure S1: Build time scales near-linearly with cohort size and benefits from thread-level parallelism. Wall-clock time for constructing the AFQuery database from single-sample VCFs (chr22) at 1, 4, 16, and 32 threads for 1,000, 5,000, and 10,000 samples. Speedup reaches 5.2× at 32 threads for 10,000 samples (24.3 s). Peak RSS: 301 MB (1,000 samples) to 1.54 GB (10,000 samples) at 32 threads.

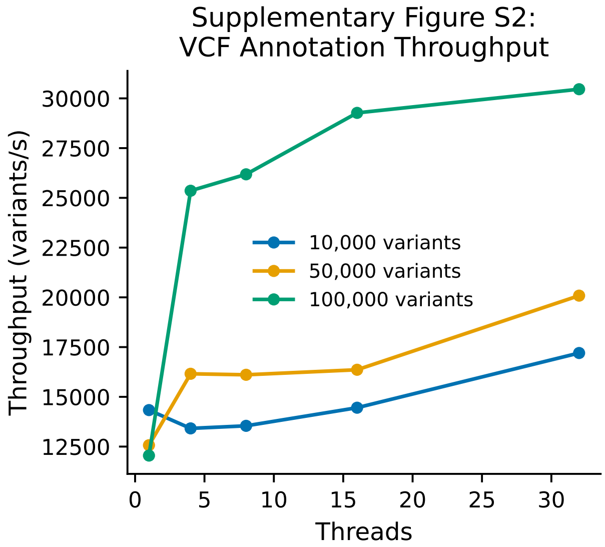

Supplementary Figure S2: VCF annotation throughput increases with variant count and thread parallelism. Throughput (variants/s) for 10,000, 50,000, and 100,000 input variants at 1, 4, and 32 threads. Maximum throughput: 30,460 variants/s at 32 threads (100,000 variants), sufficient to annotate a 30,000-variant exome in ∼1 second.

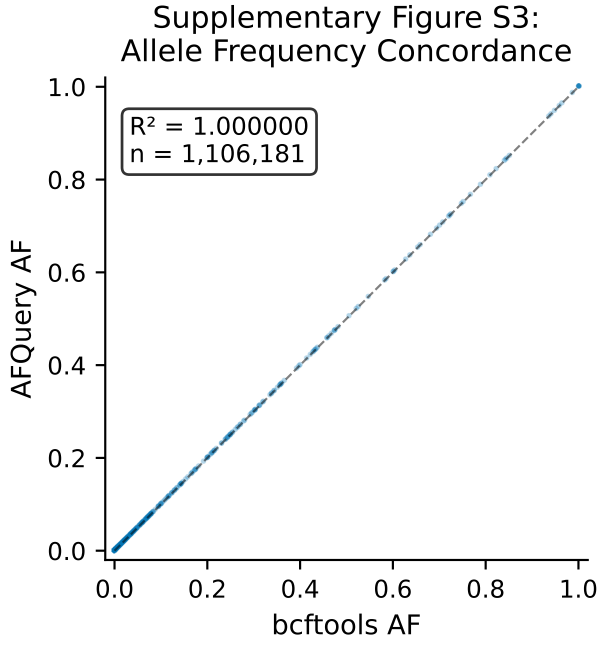

Supplementary Figure S3: AFQuery allele frequencies are concordant with BCFtools ($R^2 > 0.99999$). Scatter plot of AFQuery AF vs BCFtools AF for 1,106,181 common variants (chr22, 2,504 samples). One AC mismatch at a position with duplicate biallelic and multi-allelic records for the same alternate allele (22:17996285 A>ATCTC); AFQuery captures 8 additional carriers via per-sample ingestion ($\Delta$AC = 8). Zero AN mismatches. 4,056 variants present only in BCFtools output (all AC = 0).

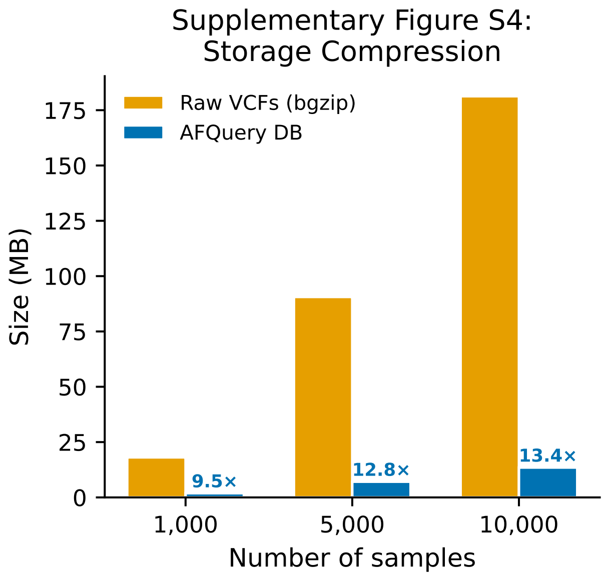

Supplementary Figure S4: Storage compression improves with cohort size. AFQuery database size vs bgzipped VCF size for chr22 at 1,000, 5,000, and 10,000 samples. Compression ratios: 9.5× (1,000 samples), 12.8× (5,000 samples), 13.4× (10,000 samples).

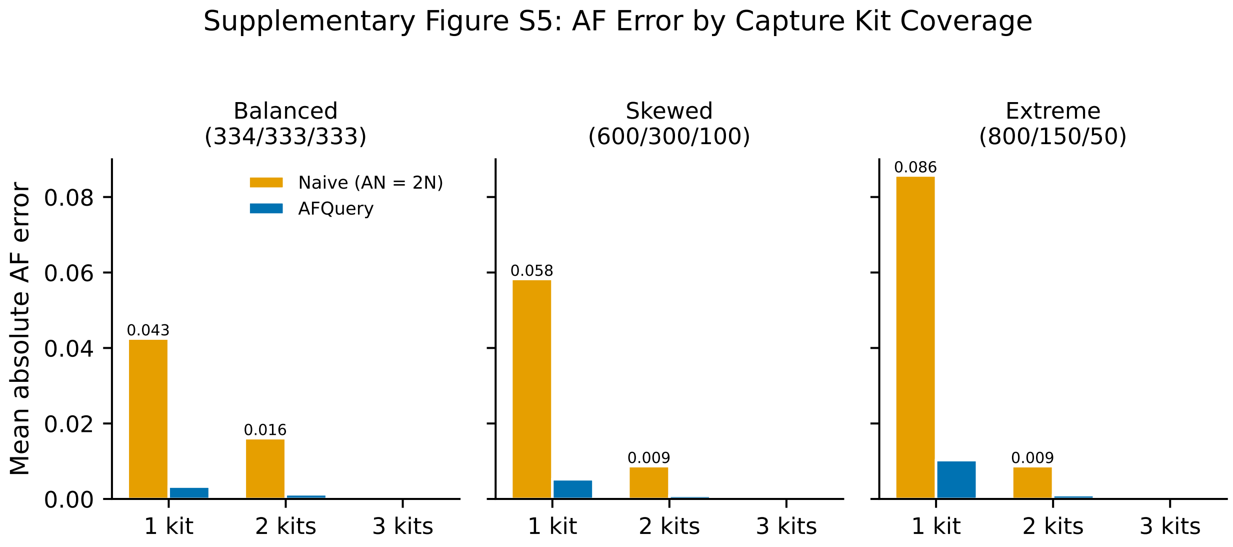

Supplementary Figure S5: AF error across all three capture kit scenarios. Extension of Figure 1B showing mean absolute error by kit coverage tier for balanced (334/333/333), skewed (600/300/100), and extreme (800/150/50) sample distributions. AFQuery MAE reduction: 13× (balanced), 11× (skewed), 8.4× (extreme). Error is concentrated exclusively at positions covered by fewer than three kits in all scenarios.

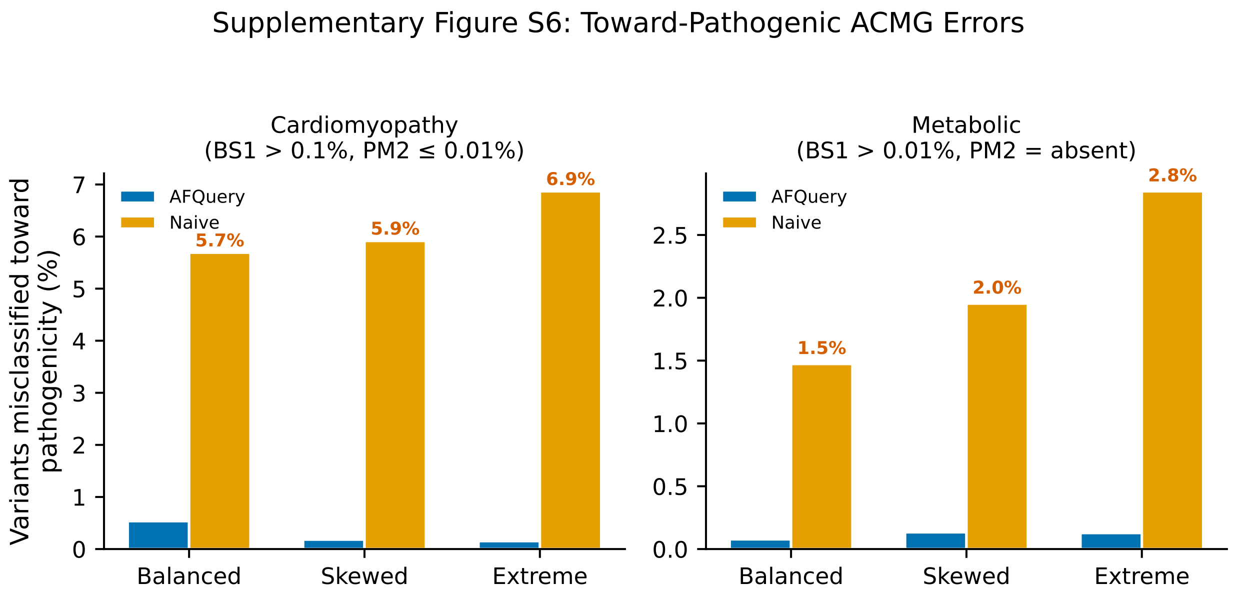

Supplementary Figure S6: ACMG classification discordance across disease types and capture kit scenarios. Toward-pathogenic error counts for AFQuery vs naive across cardiomyopathy (BA1 = 5%, BS1 = 0.1%, PM2 = 0.01%) and metabolic (BS1 = 0.01%, PM2 = absent) thresholds in all three kit scenarios. Naive errors are 100% toward pathogenicity in all six combinations (0 toward-benign errors). AFQuery toward-pathogenic errors range from 10.6× fewer (cardiomyopathy balanced) to 45.0× fewer (cardiomyopathy extreme) than naive.
